# Hyperplex PCR enables the next-generation of wastewater-based surveillance systems: long-term SARS-CoV-2 variant surveillance in Sweden as a case study

**DOI:** 10.1101/2024.06.10.24308715

**Authors:** Ruben R. G. Soares, Javier Edo Varg, Attila Szabó, Margarita Psallida, Paweł Olszewski, Danai V. Nikou, Umear Naseem, Maja Malmberg, Anna J. Székely

## Abstract

Wastewater-based epidemiology aims at measuring pathogens in wastewater as a means of deriving unbiased epidemiological information at a population scale, ranging from buildings and aircrafts to entire cities or countries. After gaining significant mainstream attention during the SARS-CoV-2 pandemic, the field holds significant promise as a continuous monitoring and early warning system tracking emerging viral variants or new pathogens with pandemic potential.

To expand the current toolbox of analytical techniques for wastewater analysis, we explored the use of Hyperplex PCR (hpPCR) to analyse SARS-CoV-2 mutations in wastewater samples collected weekly in up to 22 sites across Sweden between October 2022 and December 2023. Approximately 900 samples were tested using a dynamic probe panel with a multiplexity ranging from 10-to 18-plex, continuously adapted within 1-2 weeks to quantify relevant mutations of concern over time. The panel simultaneously covered deletions, single nucleotide substitutions, as well as variable regions resorting to probe degeneracy. By analysing all samples in parallel resorting to gold standard methods including qPCR and two different NGS technologies, the performance of hpPCR is herein shown to bridge the gap between these methods by providing (1) systematic single nucleotide sensitivity with a simple probe design, (2) high multiplexity without panel re-optimization requirements and (3) 4-5-week earlier mutation detection compared to NGS with excellent quantitative linearity and a good correlation for mutation frequency (r=0.88). Based on the demonstrated performance, the authors propose the combined use of NGS and hpPCR for routine discovery and high-frequency monitoring of key pathogens/variants as a potential alternative to the current analysis paradigm.

## 1. Introduction

Wastewater-based epidemiology (WBE), although being an 80-year-old concept, only reached the lab bench in the early 2000’s and recently catapulted to mainstream attention during the 2020 SARS-CoV-2 pandemic [1] As the name implies, the field aims at analysing wastewater samples for the presence and quantity of infectious agents to derive epidemiological information at population level. There is currently an increasing body of evidence showing the potential of this approach as a reliable and intrinsically unbiased proxy of total cases in each population, thus easing the economic and logistical burden of mass population testing [2], [3]. The latter becomes particularly relevant to enable successful implementation of pandemic preparedness systems for national level routine monitoring of circulating and emerging pathogens [4]. This approach has been tested both at city/municipality-level and at smaller scales, such as buildings (e.g. office spaces and airports) and airplanes [5], ultimately allowing to trace the origin of novel mutations to specific sites [6]. All these serve as key examples where a developed WBE toolbox can serve as a key resource in pandemic preparedness by enabling more efficient and information-driven containment policies. However, there are still multiple barriers towards reaching such degree of maturity, ranging from (1) detecting highly diluted and potentially degraded RNA in the PCR inhibitor-rich wastewater matrix, to (2) the challenges in implementing adequate sampling protocols, transport logistics and assay controls to ensure that the measured target concentrations adequately represent the population and can be reliably compared between different sampling sites and sampling times [1]. To address these challenges, sensitive, specific, mass deployable, scalable, rapid, and cost-efficient methods allowing high-frequency sampling are in demand to gather and share high volumes of data required to consolidate WBE as a routine surveillance tool.

The current analytical toolbox to measure SARS-CoV-2 RNA in wastewater comprises a first concentration and extraction-step, aiming at maximizing RNA titters while reducing the concentration of potential inhibitors [7]. Then, targeted, or non-targeted analytical methods are used for qualitative or quantitative analysis. Targeted methods include quantitative PCR (qPCR) and digital PCR (dPCR) and non-targeted analysis is typically done using next generation sequencing (NGS). Reverse transcription-qPCR (RT-qPCR) is currently the most cost-effective option and provides high sensitivity combined with a wide dynamic range, being the standard workhorse for wastewater monitoring, particularly when measuring total SARS-CoV-2 virus titters targeting conserved sequences [8]. On the other hand, despite increased running costs [9], there is an increasing shift towards ddPCR based on substantial [10], [11], albeit not undisputed [9] evidence of higher sensitivity and resistance to inhibition. However, despite significant efforts to apply both RT-qPCR [12], [13], [14], [15] and dPCR [16], [17], [18], [19] for SARS-CoV-2 variant surveillance using hydrolysis probes, protocol optimization to target point mutations typically requires extensive adjustment of annealing temperatures to avoid false-positive detections [12]. Additionally, locked nucleic acids (LNA) might have to be introduced into the probe sequence to increase specificity [15], and background signals often interfere with the results particularly for high concentrations of off-target sequences (e.g. wild-type sequence) [9]. Furthermore, both methods are limited in the degree of multiplexity per sample, often capped at less than 6 targets per sample which is suboptimal for a broad-scope variant analysis. All these factors cause delays and practical barriers in implementing fit-for-purpose assays for emerging variants, consequently forcing researchers to turn towards targeted NGS [20], [21], [22] despite the substantially higher consumable and labour costs. To address the specificity and multiplexity limitations of RT-qPCR and dPCR, while avoiding the increased costs and complexity of NGS, Hyperplex PCR (hpPCR) integrates RT-PCR, padlock probes (PLPs), rolling circle amplification (RCA) [23] and optically encoded probes [24]. This approach ensures robust single-nucleotide specificity combined with massive multiplexity of more than 100 targets per sample through post-amplification optical fluorescence imaging.

## 2. Results and Discussion

### 2.1. Methodology and study overview

The main goal of this study was to evaluate the performance of Hyperplex PCR (hpPCR) in the context of multiplex targeted quantification of SARS-CoV-2 mutations of concern in wastewater samples. The workflow of Hyperplex PCR is schematized in Figure 1-A and comprises 3 main steps. The first step includes a 1-step RT-PCR amplification of the target regions of interest, padlock probe (PLP) ligation to the target mutations and rolling circle amplification of the ligated probes. The ligation of the panel of PLPs was performed after dilution of the PCR amplicons, allowing for archiving of PCR amplicons and re-probing of mutations within the same amplicons. Performing the probing with a ligation reaction instead of typical hybridization-based hydrolysis probes (e.g. TaqMan probes or molecular beacons), particularly when using thermostable ligases ensures systematic and virtually optimization-free single nucleotide specificity under optimized conditions [25]. Each padlock probe contains a proprietary barcode in the backbone (sequence between the left and right arms) resulting in a total length of 96 bp for each probe. The subsequent RCA step generates amplification products of about 1 µm in diameter containing about 1-2 thousand copies of the barcode sequence. In the second step, the rolling circle amplification products (RCPs) are captured on a surface and each barcode will be specifically recognized by Nanopixel probes having a unique optical code corresponding to each target barcode of the PLPs. In the final third step, the surface is imaged with fluorescence microscopy and RCPs of each specific optical barcode are counted and normalized to an internal RCP reference to neutralize variation in capture efficiency.

**Figure 1:**
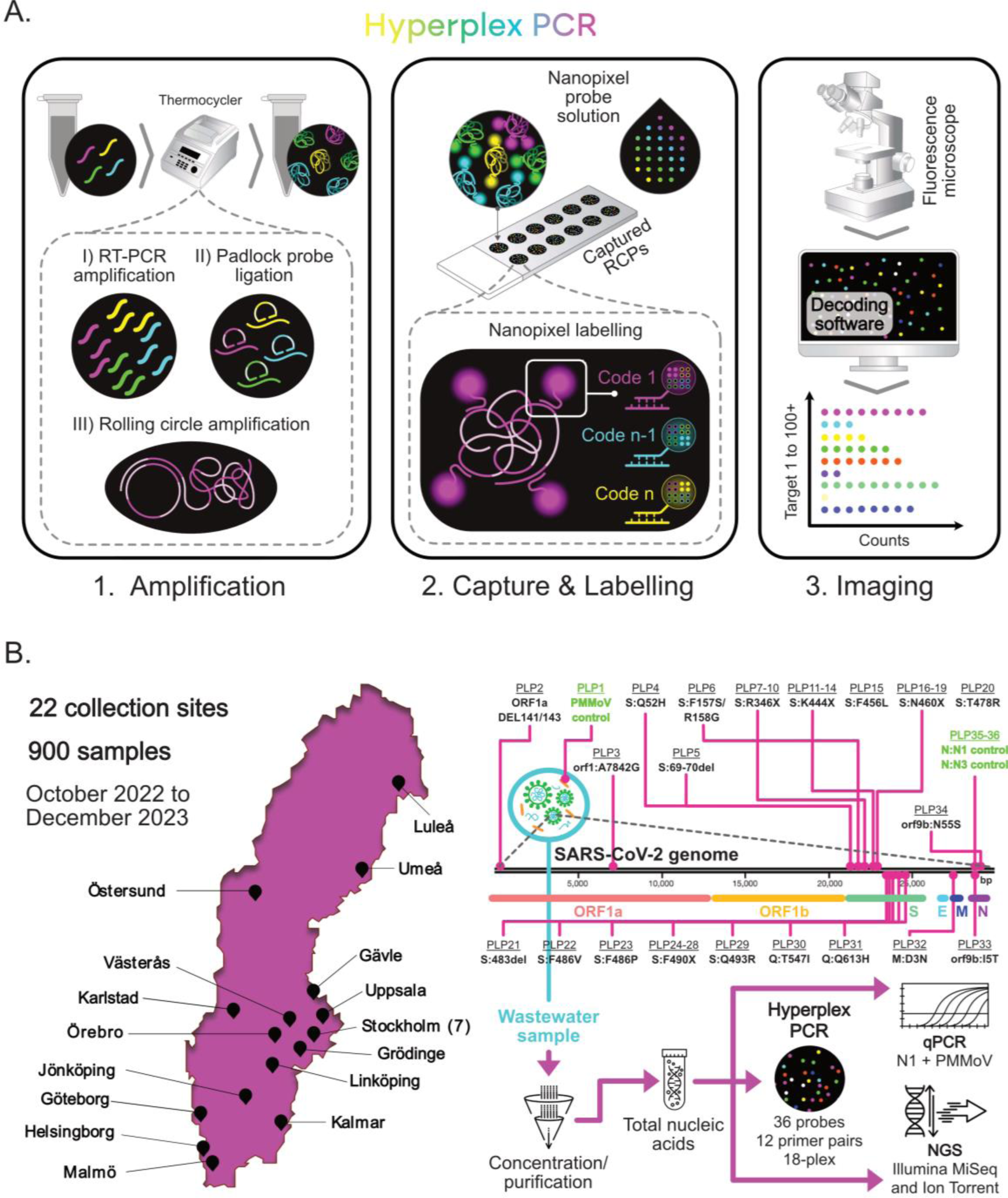
Hyperplex PCR workflow and overview of wastewater collection and analysis approach. **A-** Workflow of Hyperplex PCR comprising 3 main analytical steps: (1) amplification of extracted nucleic acids by reverse transcription PCR (RT-PCR), followed by padlock probe (PLP) ligation and re-amplification of the PCR amplicons into rolling circle amplification products (RCPs); (2) Capture of the RCPs on a solid phase with a SLAS format and subsequent hybridization to Nanopixel probes and; (3) Imaging of the Nanopixel-probed RCPs using an epifluorescence microscope, followed by decoding of the Nanopixel optical probes using a proprietary image analysis tool from APLEX Bio which converts the image into RCP counts per PLP target sequence. **B-** Design of long-term study and analytical approach using Hyperplex PCR, qPCR, Illumina and Ion Torrent in parallel for each extracted sample. For Hyperplex PCR, a total of 12 PCR primer pairs and 36 PLPs were designed spanning the entire genome of SARS-CoV-2. To adapt to outcompeted and emerging viral variants over time, not all probes were used simultaneously and up to 18-plex target analysis was performed. The “X” in the amino-acid substitution nomenclature refers to multiple mutations being covered by a set of degenerate PLPs encoded by the same Nanopixel probe

Figure 1-B illustrates the collection sites, probe design map within the SARS-CoV-2 genome, and sample analysis logistics. From October 2022 to December 2023, approximately 900 wastewater samples were collected weekly from up to 22 wastewater treatment plants across Sweden, including 6 located in Stockholm municipality (Table S7). During the analysis period a total of 36 PLPs were designed targeting 12 RT-PCR amplified regions of the SARS-CoV-2 genome. Three probes were dedicated to internal control targets, namely PMMoV as a marker of human fecal material and N1 (up to w15/2023) or N3 (from w16/2023) as conserved SARS-CoV-2 genes. The remaining 33 probes were designed to target key mutations characteristic of emerging SARS-CoV-2 variants of concern. Among this probe set, (1) single nucleotide substitutions, (2) deletions and (3) pooled mutation profiling were achieved simultaneously with different probe designs. PLPs for single nucleotide substitutions were preferably designed with the mutation placed on the 3’ end, PLPs for deletions were designed with the extremes of the deleted region complementary to each probe arm and pooled mutation profiling (designated as “X” amino acid substitutions in Figure 1-B) was achieved using degenerate bases and common barcodes (corresponding to the same Nanopixel probe) in the PLP backbone. The PCR primers and PLPs included in the analysis were modified throughout the study (Methods section 4.4.1), from initially 10-plex to an 18-plex panel by Summer 2023 upon the emergence of the BA.2.86 variant.

Since one of the key objectives of the study was to evaluate if Hyperplex PCR could improve WBE analysis compared to current methodologies, in particular non-targeted approaches, all collected samples were split after extraction and measured in parallel using conventional qPCR and one of two NGS methods, namely optical-(Illumina) and semiconductor-based (Ion Torrent) sequencing. (Ion Torrent: w40/2022-w21/2023 and w35-36/2023, and Illumina: w22-34/2023 and w37-52/2023). The following subsections explore the output of Hyperplex PCR compared to qPCR, and NGS, as well as published clinical data to validate its relevance in the context of WBE and future pandemic preparedness.

### 2.2. Hyperplex PCR allows multiplexed relative quantification of at least 18 mutations of concern per assay with rapid panel customization

The qPCR and hpPCR results obtained for all tested samples are summarized as heat maps in Figure 2. For qPCR the color coding corresponds to concentration of N1 copies normalized to PMMoV. For hpPCR, the results are expressed as relative counts, meaning the absolute signal of the assay for a specific target (according to methods section 4.4.3) normalized to the highest absolute signal measured for that same specific target within the entire dataset. This relative normalization was applied since the efficiency of PCR amplification was not calibrated between targets, hence making the absolute counts not comparable between different mutations, particularly those targeting sequences in different PCR amplicons, but comparable between different time-points (x-axis) and collection sites (y-axis). While the assay could be designed to provide absolute quantification for each mutation with appropriate internal controls and/or calibration curves, the original aim of the application of the hpPCR method was to complement the ongoing viral quantification efforts with variant composition assessment. Overall, the results in Figure 2 highlight the expanded dataset obtained using hpPCR compared to qPCR, allowing the discrimination of trends for several key mutations of concern according to the recommendations of health authorities at a given time point. Since PLPs ensure reliable single nucleotide specificity with minimal requirements of assay optimization, the targets in the panel can be adapted on demand by including new primers/probes for emerging mutations of concern (solid vertical lines in Figure 2) or excluding primers/probes targeting outcompeted mutations (dashed vertical lines in Figure 2). Upon emergence of a new mutation of interest, the delay to implement a new probe was less than 2 weeks including design (< 1 day), oligo procurement (5-10 days), and PCR primer/PLP mix formulation (< 1 day). Two main waves of infection were observed throughout the testing period, during the winter season of 2022 and autumn/winter season of 2023. During these periods of increased SARS-CoV-2 viral titters in wastewater samples, already prevalent and particularly emerging mutations were also found to peak during those periods, namely S:346X (XBB, BA.2.75), S:K444X (BQ.1), S:N460X (BQ.1, BA.2.75, XBB) and S:F490S (BJ.1, XBB.1) during the first wave and BA.2.86 mutations (S:483del, S:F157S/R158G, Orf1:A7842G and S:69-70del) during the second wave. Overall, the trends measured for each mutation provided an excellent correlation with trends observed for clinical cases during periods of active clinical monitoring (Figure S1, where the clinical cases for each mutation where estimated using the CoV-Spectrum Portal [26] and the total SARS-CoV-2 incidence published by The Public Health Agency of Sweden - data accessed on March 2023) and successfully allowed the surveillance of re-emerging mutations such as S:K444T from the first clinical cases of DV.7.1 in w30/2023 (asterisk in Figure 2) until virtually zero incidence by w52/2023. Clinical incidence of DV.7.1 was obtained from the CoV-Spectrum portal [26] accessed in March 2024. Considering that upon the decision to implement probes targeting S:F456L, S:Q52H and BA.86 mutations, positive signals were found in most of the tested cities, a retrospective analysis was performed for the 8 sites covering more than 100.000 people in the catchment area back to w16/2023 to test the early detection of these variants. The results of this retrospective analysis are described below in section 2.3.

**Figure 2:**
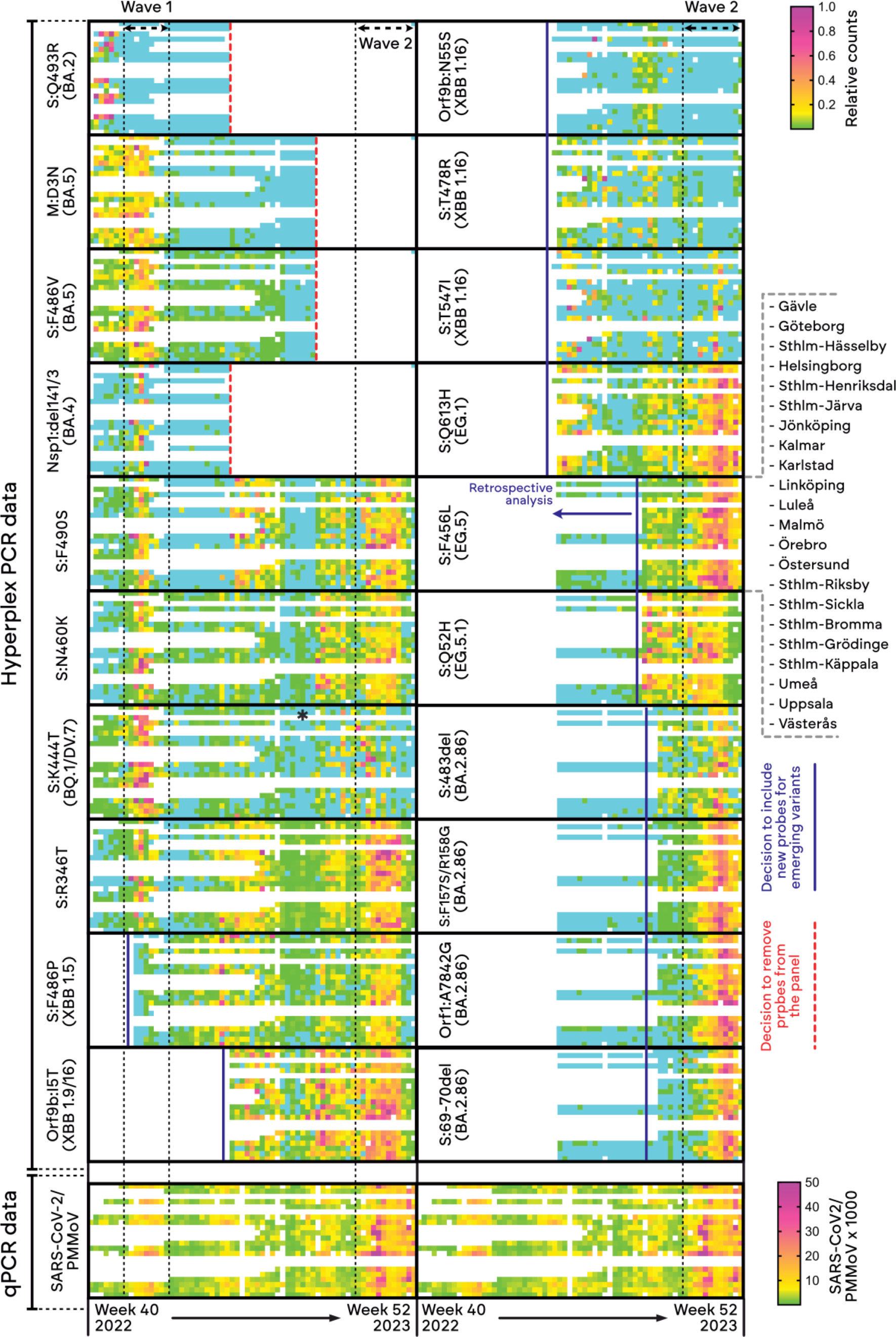
Heatmap plot summarizing the Hyperplex PCR and qPCR results obtained for all tested samples between October 2022 and December 2023 with a frequency of one sample per week for each collection site. qPCR targeted SARS-CoV-2 using the N1 assay until w29/2023 and SCV2 onwards. For each target sequence, columns correspond to weekly measurements and rows to each of the 22 collection sites (Table S7). For Hyperplex PCR relative counts refer to signal obtained for each target sequence normalized to the maximum intensity measured for each target among all samples. The blue color code refers to signals below the detection threshold. Variants indicated for each mutation refer to the most prevalent variant associated to the mutation at the time the probes were included in the panel. The asterisk in the S:K444T panel refers to the identification of the first clinical cases of DV.7.1 in Sweden. The qPCR data is duplicated below each column of 10 hpPCR targets for visualization purposes.

### 2.3. Hyperplex PCR provides mutation frequency quantification comparable to NGS with significantly higher sensitivity

Expanding on the quantitative capabilities of hpPCR beyond the relative quantification of target mutations over time in multiple sites, additional probes targeting the wild-type sequence of specific target mutations were added to evaluate the quantification of mutation frequency (MF). By quantifying MF it becomes possible to compare the prevalence of different target mutations by having the total virus (Wt plus Mut sequences) as internal control. In this case, two gold standard NGS methods typically used for wastewater analysis, namely Illumina and Ion Torrent were used to benchmark the MF output of the hpPCR assay focusing on samples collected around the period of wave 1. The MF quantification concept and summary of results are shown in Figure 3-A. According to the example schematics and imaging example of 2-plex results for mutation S:R346T, two uniquely barcoded PLPs are used to target the wild-type (G22599) or mutated sequences (G22599C). The plots highlight the following three key observations. (1) A good correlation of hpPCR and NGS when measuring MF over time for each of the four target mutations was observed, here tested for 2 different major populational centers, namely Uppsala and Malmö covering 191 000 and 356 000 people in the catchment area, respectively. Importantly, both measurements agree with the proportion of SARS-CoV-2 clinical samples found with these mutations in Sweden during this period (Figure S2). (2) Spiking known mixtures of synthetic amplicons containing each of the target mutations, hpPCR provides an excellent linearity (R^2^=0.99) and agreement between expected and observed MF values. (3) Pooling all data obtained for both Uppsala and Malmö samples between w42/2022 and w48/2022, a Pearson correlation coefficient of 0.88 was obtained between the MF output of hpPCR and NGS. Overall, these observations indicate that hpPCR can serve as a viable alternative to NGS for the targeted quantification of point mutation frequency in wastewater. Furthermore, hpPCR was further compared with NGS concerning sensitivity to allow early detection of emerging mutations of concern according to the results in Figure 3-B. Using hpPCR it was systematically possible to detect positivity for emerging mutations 4 to 5 weeks earlier compared to NGS for 3 key mutations characteristic of highly relevant VOCs, namely XBB 1.5 and EG.5 and a set of mutations characteristic of BA.2.86. Remarkably, the readout using hpPCR provided a superior correlation with clinical incidence and early detection when mutation specific incidence represented less than 2% of the total SARS-CoV-2 clinical cases. In the case of BA.2.86, which had several differentiating characteristic mutations relative to other coexisting XBB and BA.5-descendant variants, based on early sequencing data from Swedish patients, only a small subset of 4 mutations were selected for monitoring using hpPCR. Nevertheless, even considering all BA.2.86 characteristic mutations measured using NGS, hpPCR still provided a 3-week earlier detection targeting 4 mutations within the same list of BA2.86 characteristic mutations considered for NGS positivity (Figure S3, Table S6). Interestingly, two samples with at least 3 key mutations of BA.2.86 were found in w20 and w21, in line with the results of NGS considering all BA.2.86 mutations, albeit for different targets and sites (Figure S3, Table S6). Ultimately, combining the possibility of rapidly implementing probes with NGS-grade specificity but having a sensitivity significantly higher than NGS, hpPCR intrinsically enables earlier detection of targets than currently available targeted and non-targeted gold standard methods.

**Figure 3:**
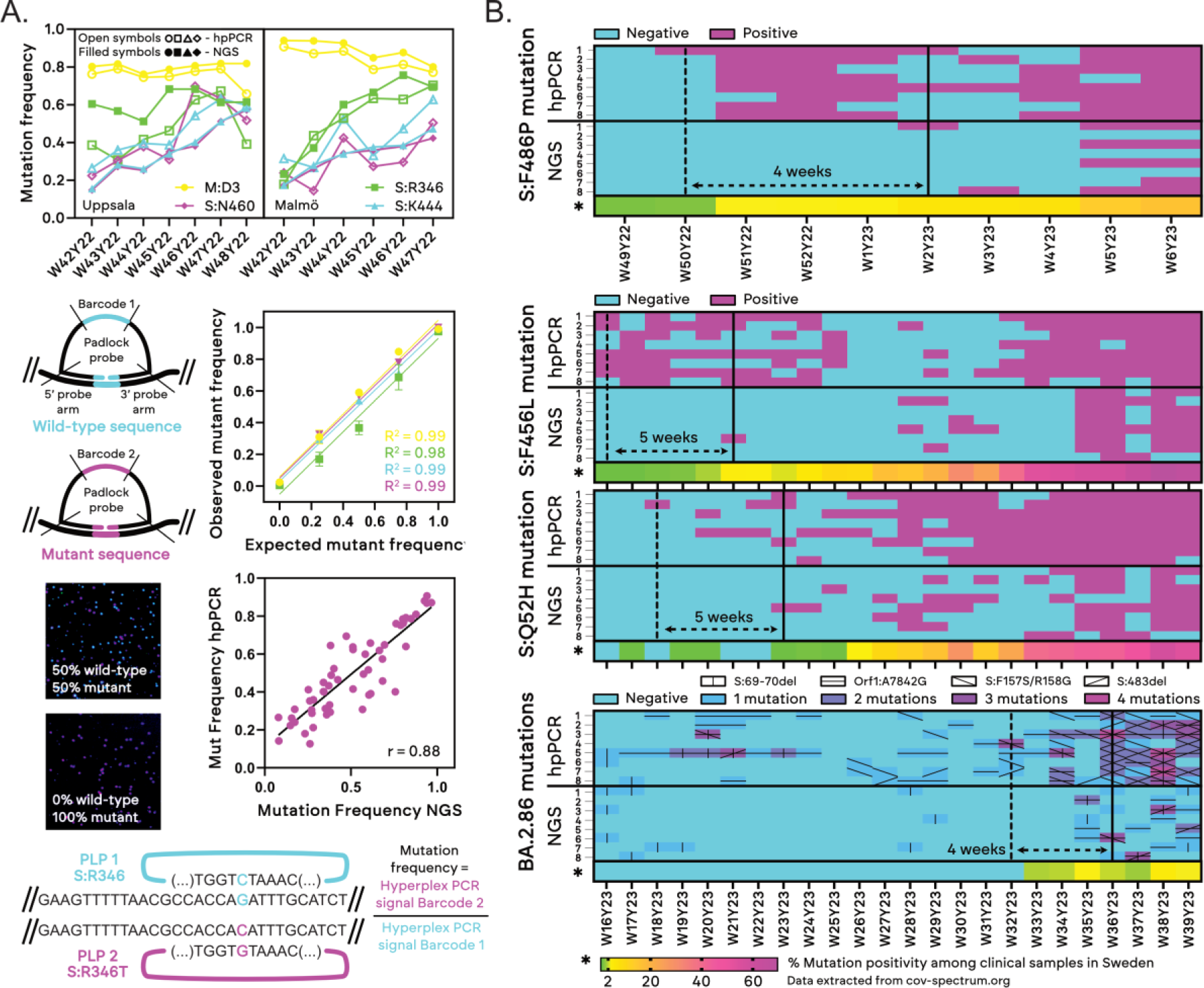
Benchmarking of quantitative performance (A) and sensitivity (B) of Hyperplex PCR against next-generation sequencing (NGS). **A-** Conceptual schematic and demonstration of Hyperplex PCR used to quantify mutation frequency of target single nucleotide substitutions, namely M:D3N (G26529A), S:R346T (G22599C), S:N460K (T22942G) and S:K4444T (A22893C). Two probes with different Nanopixel barcodes were used for each target mutation, one targeting the wild-type sequence and the other the mutated sequence. For Hyperplex PCR, the mutation frequency is calculated as the ratio of counts (RCPs) for the mutant sequence to those of the wild-type sequence. The plot showing expected vs observed mutant frequency was obtained using synthetic PCR amplicons (ssDNA) spiked in solution. The plot showing correlation between hpPCR and NGS mutation frequency was obtained by testing wastewater samples (w42/2022 to w48/2022) targeting M:D3, S:N460, S:R346 and S:K444. **B-** Early detection of mutations and variants of concern using Hyperplex PCR vs NGS. S:F486P, S:F456L and S:Q52H were characteristic of variants XBB 1.5, EG.5 and EG.5.1 when measurements were initiated. Tested mutations for BA.2.86 include S:483del, S:F157S/R158G, Orf1:A7842G and S:69-70del. NGS data refers to the same mutations targeted by Hyperplex PCR. The y axis refers to different collection sites as follows: 1-Göteborg, 2-Helsingborg, 3-Malmö, 4-Örebro, 5-Stockholm-Käppala, 6-Umeå, 7-Uppsala, 8-Västerås. The heatmap below each chart refers to the fraction of clinical samples collected in Sweden found to be positive for each of the target mutations under analysis.

## 3. Conclusions

Despite the mounting evidence of the potential of WBE for surveillance of present and future pathogen outbreaks and pandemics, the field is still in its infancy with regards to method and output standardization. The current typical WBE approach is schematized in Figure 4 according to alternative 1 where surveillance typically focuses on quantitative monitoring of a limited number of pathogens of interest using target specific qPCR or dPCR. For the surveillance of variants and mutations, this is complemented either by NGS, which is typically successful only in case of sufficiently high titers, or alternatively, by individual mutation specific qPCR or dPCR assays. In this approach there is often a dilemma between reagent and labor cost-effectiveness and necessary performance including the optimization of qPCR or dPCR assays for novel targets, which often culminates with researchers circling back to costly NGS, particularly when several point mutations, become targets of interest as is the case for most SARS-CoV-2 viral variants. Alternatively, Hyperplex PCR is herein demonstrated as a powerful tool enabling affordable quantitative and qualitative high-frequency multi-pathogen wastewater monitoring, deployable in virtually any research lab. As demonstrated here has a performance competitive with state-of-the-art NGS methods, the possibility of rapidly including new probes on-demand and a simple workflow with results from extracted RNA within one workday. Towards enabling a next-generation single-method surveillance approach, Hyperplex PCR thus successfully fills the bioanalytical method gap between qPCR/dPCR and NGS, upgrading the current WBE analytical toolbox by providing: (1) High multiplexity per sample with single nucleotide specificity, (2) NGS-grade mutation frequency quantification with a good correlation (r = 0.88) extremely high linearity (R^2^ > 0.99), (3) 4-weeks+ early detection of variants of concern demonstrated vs sequencing, (4) Panel customization with new probes within 2 weeks without panel re-optimization. Based on the results of the long-term study reported herein, the authors believe that hpPCR, particularly upon maximization of multiplexity towards the theoretical limits of 100+ target sequences and complemented with open data platforms for rapid probe design and panel customization has the potential to become a gold-standard for sustainable and standardized WBE.

**Figure 4:**
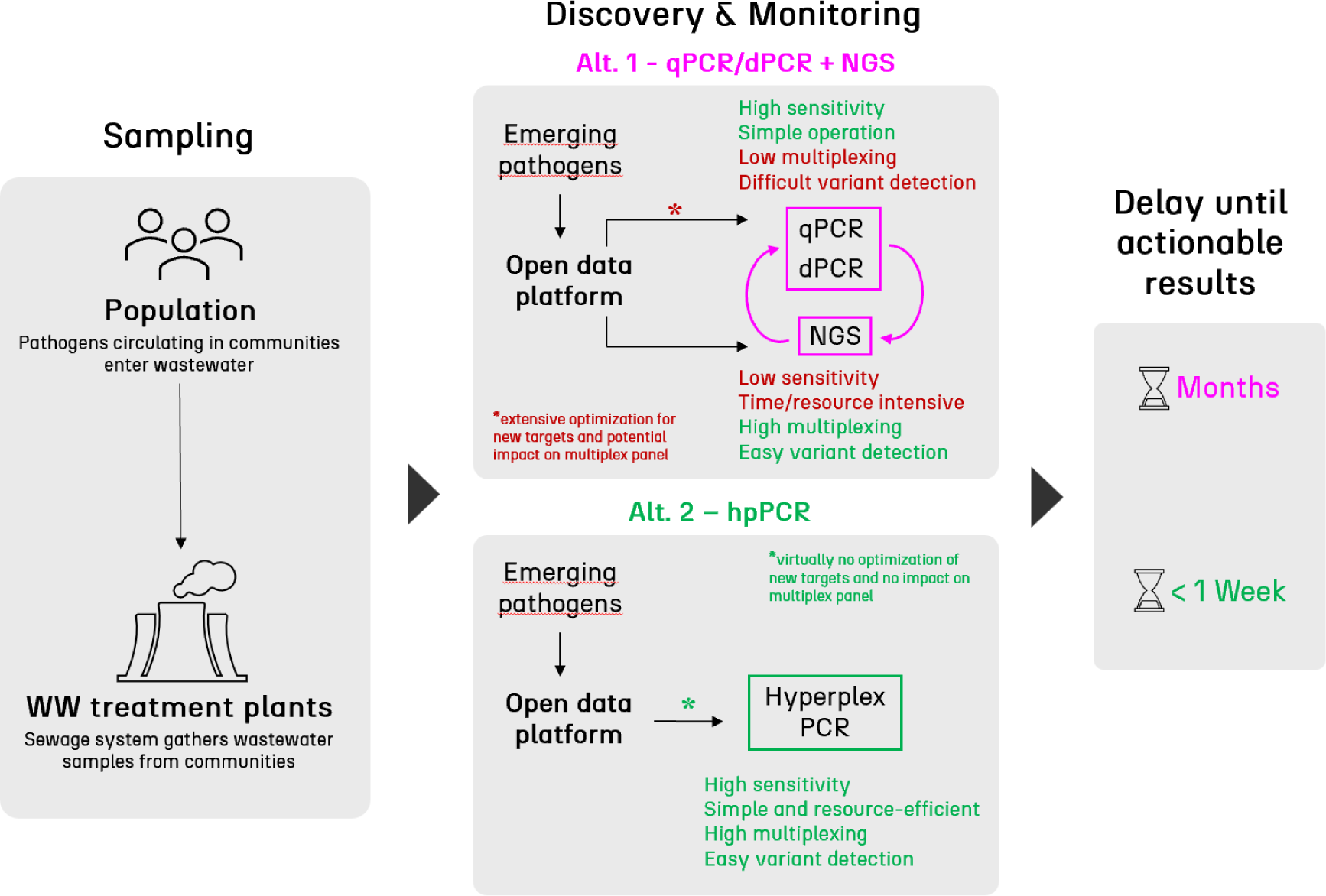
Current approach typically used for WBE during and after the SARS-CoV-2 pandemic (Alt.1) resorting to qPCR, dPCR and NGS for high-frequency monitoring and proposed alternative approach (Alt. 2) using only hpPCR. The delay until actionable results includes the time to design and validate new primers/probes for emerging mutations/variants in the case of qPCR/dPCR and/or the time to select the ideal method among qPCR, dPCR and NGS to strike an adequate balance between sensitivity and specificity of mutation identification within a certain setting.

Figure 1: Hyperplex PCR workflow and overview of wastewater collection and analysis approach. **A-** Workflow of Hyperplex PCR comprising 3 main analytical steps: (1) amplification of extracted nucleic acids by reverse transcription PCR (RT-PCR), followed by padlock probe (PLP) ligation and re-amplification of the PCR amplicons into rolling circle amplification products (RCPs); (2) Capture of the RCPs on a solid phase with a SLAS format and subsequent hybridization to Nanopixel probes and; (3) Imaging of the Nanopixel-probed RCPs using an epifluorescence microscope, followed by decoding of the Nanopixel optical probes using a proprietary image analysis tool from APLEX Bio which converts the image into RCP counts per PLP target sequence. **B-** Design of long-term study and analytical approach using Hyperplex PCR, qPCR, Illumina and Ion Torrent in parallel for each extracted sample. For Hyperplex PCR, a total of 12 PCR primer pairs and 36 PLPs were designed spanning the entire genome of SARS-CoV-2. To adapt to outcompeted and emerging viral variants over time, not all probes were used simultaneously and up to 18-plex target analysis was performed. The “X” in the amino-acid substitution nomenclature refers to multiple mutations being covered by a set of degenerate PLPs encoded by the same Nanopixel probe.

## 4. Methods

### 4.1. Wastewater sample collection and extraction

For each site, we collected weekly samples of untreated wastewater integrated over a single day (24 hours) using flow-compensated samplers. All measurements represent only one day except for Uppsala, where daily samples were combined flow-proportionally into one composite sample representing one week. The processing of samples followed the protocols outlined in Isaksson et al. [27] involving the concentration and extraction of viral genomic material using the Maxwell RSC Enviro TNA kit (Promega). All nucleic acid extracts were stored at -80 °C until further processing.

### 4.2. qPCR quantification of SARS-CoV-2 and PMMoV

The absolute quantification of SARS-CoV-2 genome copy numbers followed the methodology outlined in Isaksson et al. [27]. Until w31/2023 quantification was performed using the SARS-CoV-2 specific N1 assay [28] while from w32/2023 the Flu SC2 Multiplex Assay (CDC) was utilized. To account for variations in population size and wastewater flow, the SARS-CoV-2 genome copy numbers were normalized to pepper mild mottle virus (PMMoV) copy numbers. PMMoV quantification followed a modified version of the assay described by Zhang et al [29]

### 4.3. Next generation sequencing (NGS)

The generated total nucleic acid (TNA) extracts were sequenced either on Thermo Fisher’s Ion Torrent System (w40/2022-w21/2023 and w35-36/2023), or on Illumina (w22-34/2023 and w37-52/2023).

#### 4.3.1. Ion torrent sequencing

The sequencing libraries for Ion Torrent were prepared from the extracted total nucleic acids were prepared using the Ion AmpliSeq™ SARS-COV-2 Insight Research Assay (Thermo Fisher Scientific) and the SuperScript™ VILO™ cDNA Synthesis Kit (Thermo Fisher), following the manufacturer’s protocols “Reverse transcribe RNA with the SuperScript™ VILO™ cDNA Synthesis Kit” (Thermo Fisher) and “Prepare libraries on the Ion Chef™ Instrument” (Thermo Fisher) with the Ion Chef System. Sequencing was conducted on the Ion S5 XL System (Thermo Fisher) by multiplexing using Torrent Suite Software version 5.16.1 as per to the manufacturer’s instructions (Thermo Fisher). The generated sequence data were filtered, aligned to the manufacturer’s references (Ion_AmpliSeq_SARS-CoV-2-Insight_Reference), and used to generate mutation frequency tables for variant detection using the manufacturer’s analysis pipeline in Torrent Suite 5.16 (Thermo Fisher, # MAN0017972 Rev B.0, Ch 12). Library preparation, Ion Torrent sequencing, and sequence analyses were carried out at the National Genomics Infrastructure (NGI) at Science for Life Laboratory, Uppsala, Sweden.

#### 4.3.1. Illumina sequencing

For the Illumina sequencing, DNAse treatment (DNase I, Amplification Grade, Invitrogen) was performed to reduce unspecific amplification before library preparation by the COVIDSeq Assay kit (Illumina). Samples from weeks 22, 23, 33, and 34/2023 were amplified using the Artic V4.1 NCOV-2019 Panel primers (IDT DNA), while the remaining samples sequenced by Illumina technology were amplified using the Artic V5.3.2 NCOV-2019 Panel primers (IDT DNA). The resulting libraries were sequenced either on the Illumina MiSeq platform using the MiSeq Reagent Kit v3 2 x 76 (150-cycle) (Illumina) (w22-34/2023 and w40/2023), or the Illumina NextSeq 550 platform using the NextSeq Reagent Kit 500/550 High Output Kit v2 2 x 74 (150-cycle) (Illumina) (w37-39/2023 and w41-52/2023). The generated sequence data were processed and used to generate mutation frequency tables for variant detection using variant quantification in sewage pipeline designed for robustness (VaQuERo.v2) [2]. Library preparation, Illumina sequencing, and sequence analyses were carried at the Swedish University of Agriculture Sciences (SLU).

### 4.4. Single-reaction 18-plex mutation detection with Hyperplex PCR

The Hyperplex PCR reaction comprises 3 main steps, (1) PCR amplification, PLP ligation and RCA; (2) RCP capture and probing with Nanopixel probes; (3) Imaging of the surface using a fluorescence microscope. The amplification, capture and labelling protocol was performed according to instructions of a custom Hyperplex PCR kit assembled by APLEX Bio including the PCR primers and PLP sequences listed in Table 1.

#### 4.4.1. Probe design, PCR amplification, PLP ligation and RCA

To maximize PCR efficiency, all PCR primers according to Table S1 were designed to generate amplicons with less than 250 bp and having within 5% CV of melting temperature averaging 63 °C. Reference sequences for each target mutation and/or variant were obtained via the European COVID-19 Data Portal and the GISAID initiative [30]. Depending on sequence availability at the date probes were included in the panel, 15 to 200 sequences from Swedish patients were aligned and used for primer and probe design. RT-PCR amplification was performed by combining 2 µL of extracted nucleic acids with 8 µL a master mix containing all relevant primers for the targets under analysis. Final primer concentrations were between 50 and 150 nM for the forward primers and 67 to 300 nM for the reverse primers according to Tables S2, S3, S4 and S5. The mix was then subjected to the following temperatures using a thermal cycler: 25°C for 2 min, 50°C for 15 min, 95°C for 2 min, followed by 40 cycles of [95°C for 3 sec, 60°C for 1 min]. After PCR, the amplicons were diluted with 190 µL DNase/RNase-free water water and 0.5 µL of the diluted mixture were combined with 19.5 µL of Ligation mixture in a separate PCR tube, comprising the mix of PLPs (50 pM to 1 nM each final concentration), ligation buffer and hpPCR Ligase. For PLP ligation, the mixture was placed in the thermocycler and subjected to 95 °C for 20 s and 60 °C for 30 min. 10 µL of RCA master mix containing hpPCR polymerase were then added to the 20 µL ligation mix and subjected to an additional 2 hours at 37 °C, followed by enzyme inactivation at 60 °C for 20 min. All temperature cycling and incubation steps were performed in a Bio-Rad T100 thermal cycler with default temperature ramp conditions.

#### 4.4.2. RCP capture and probing with Nanopixels

After RCA, 2.5 µL of the RCP solution was combined with 50 µL of capture buffer containing an RCP reference. The RCP reference serves as an internal control to normalize the capture efficiency of RCPs on the surface. 40 µL of this mix were then transferred into the wells of the slide frame mounted on the capture slide provided in the kit, followed by incubation for 5 min at room temperature. Afterwards, 100 µL of absolute EtOH were added to each well on top of the capture solution and all contents were subsequently discarded. The wells were then washed twice with wash buffer, followed by 50 µL of blocker buffer which was left to incubate 5 min at room temperature. The solution was then removed and 40 µL of labelling master mix containing the probes were added. After 60 min incubation at 37 °C using an incubator oven (Memmert BE200), the solution was removed, each well washed four times with 50 µL wash buffer, the slide frame was disassembled and finally mounted using antifade solution (SlowFade™ Diamond Antifade Mountant, Thermo Fisher Scientific).

#### 4.4.3. Fluorescence microscopy imaging

The microscope used for imaging was a Zeiss Axio Imager 2 equipped with a Hamamatsu Orca Fusion sCMOS camera, a Colibri 7 solid-state light source, a 20x Plan-Apochromat 20x/0.8 M27 objective and the following emission filter sets from CHROMA: 525/50 BP, 605/52 BP, 690/50 BP, 785/25 BP. Image acquisition was performed with 9 tiles per sample. Analysis of the raw czi files was performed using a proprietary software tool from APLEX Bio including the following general functions: (1) image cropping and background subtraction, (2) channel alignment, (3) segmentation, (4) spot processing and extraction, (5) Nanopixel decoding algorithm and (6) output of counts per color code normalized to an internal reference. For all target mutations, the signal was corrected (cutoff baseline) using an internal control sample containing 1.000 copies/μL of wild-type SARS-CoV-2 IVT RNA (Twist Bioscience Viral Control 2, Wuhan-Hu-1, MN908947.3). All signals above the detection threshold are higher than the average signal of the control plus 3.29 × SD (n=4).

## Data Availability

All data produced in the present study are available upon reasonable request to the authors

## Acknowledgements

The data handling was enabled by resources provided by the National Academic Infrastructure for Supercomputing in Sweden (NAISS), partially funded by the Swedish Research Council through grant agreement no. 2022-06725. Ion Torrent sequencing was performed by the SNP&SEQ Technology Platform in Uppsala. The facility is part of the National Genomics Infrastructure (NGI) Sweden and Science for Life Laboratory. The SNP&SEQ Platform is also supported by the Swedish Research Council and the Knut and Alice Wallenberg Foundation. This work was funded by governmental grants provided to the Public Health Agency of Sweden for assignment S2022/04841. We further acknowledge the Public Health Agency of Sweden for contributions to padlock probe target design. U.N. and R.S. are co-inventors of the hpPCR methodology according to patent application WO/2021/206614. U.N. holds shares in Aplex Bio AB. Other authors declare no competing financial interests.

