## Supplementary information file for "Hyperplex PCR enables the next-generation of wastewater-based surveillance systems: long-term SARS-CoV-2 variant surveillance in Sweden as a case study"

[*] Contact authors:

Ruben R.G. Soares; Nobels väg 16, Solna, Sweden;

Anna J. Székely; Lennart Hjelms väg 9, Uppsala, Sweden;

**Table S1:** Sequences of PCR primers and padlock probe arms used for long-term relative quantification of SARS-CoV-2 mutations in wastewater samples according to the results in Figure 2. “p” refers to 5’-phosphorylation.

| Sequence type | Name | PLP arm | Sequence (5'-3') |
|---|---|---|---|
| Padlock probes | N1 | left | pCGTTTGGTGGRCCCTCAGAT |
| | | right | GCGAAATGCACYYCGCATTA |
| | N3 | left | pCCGCAATCCTGCTAACAATG |
| | | right | CCAAAAGATCACATTGGCAC |
| | PMMoV | left | pAAGCAAATGTCGCACTTGCA |
| | | right | AACGTTTGAGAGGCCTACCG |
| | M:D3N | left | pATTCCAACGGTACTATTACC |
| | | right | TTTAATTTTAGCCATGGCAA |
| | S:F486V | left | pTTAATTGTTACTTTCCTTTA |
| | | right | TTGTAATGGTGTTGCAGGTG |
| | S:R346T | left | pATTTGCATCTGTTTATGCTT |
| | | right | GAAGTTTTTAACGCCACCAC |
| | S:R346S | left | pYTTTGCATCTGTTTATGCTT |
| | | right | GAAGTTTTTAACGCCACCAG |
| | S:R346X | left | pATTTGCATCTGTTTATGCTT |
| | | right | GAAGTTTTTAACGCCACCRW |
| | S:N460K | left | pTTAGAYTTCCTAAAMAATCT |
| | | right | CTCTCTCAAAAGGTTTGAGY |
| | S:N460X | left | pTAGACTTCCTAAACAATCTA |
| | | right | TCTCTCAAAAGGTTTGAGAM |
| | S:K444T | left | pGGTTGGTGGTAATTATAATT |
| | | right | TCTAACAAGCTTGATTCTAC |
| | S:K444N | left | pYGTTGGTGGTAATTATAATT |
| | | right | TCTAACAAGCTTGATTCTAA |
| | S:K444X | left | pGGTTGGTGGTAATTATRATT |
| | | right | TCTAACAAGCTTGATTCTAK |
| | S:F490S(1) | left | pTCCTTTACRATCATATGGTT |
| | | right | GMAGGTKYTAATTGTTACTC |
| | S:F490S(2) | left | pTCCTTTACAATCATATGGTT |
| | | right | GCAGGTCCTAATTGTTACTC |
| | S:F490V | left | pTTCCTTTACRATCATATGGT |
| | | right | TGCAGGTTTTAATTGTTACG |
| | S:F490X | left | pTCCTTTACRATCATATGGTT |
| | | right | GMAGGTTTTAATTGTTACYR |
| | nsp1:del141/143 | left | pTAGATCGGCGCCGTACCTAT |
| | | right | CCAAGCTCGTCGCCTAAGTC |
| | S:Q493R | left | pGATCATATGGTTTCCGACCC |
| | | right | TAATTGTTACTTTCCTTTAC |
| | S:F486P | left | pCTAATTGTTACTYTCCTTTA |
| | | right | TTGTAATGGTGTTGCAGGTC |
| | S:T478R | left | pACCTTGTAATGGTGTTGCAG |
| | | right | ATCTATCAGGCCGGTAACAG |
| | S:T547I | left | pTTAAACCATTGAAGTTGAAA |

| | | | |
|---|---|---|---|
| | | right | AGTAAGAACACCTGTGCCTA |
| | orf9b:I5T | left | pCAGCGAAATGCACTCCGCAT |
| | | right | CTGATAATGGACCCCAAAAC |
| | orf9b:N55S | left | pCATGGCAAGGAAGACCTTAA |
| | | right | GGTTCACCGCTCTCACTCAG |
| | S:Q613H | left | pGGTGTTAACTGCACAGAAGT |
| | | right | AGGTTGCTGTTCTTTATCAT |
| | S:F456L | left | pAACAATCTATACAGGTAAT |
| | | right | TTGAGCTTAGACTTCCTT |
| | S:Q52H | left | pGACTTGTTCTTACCTTTCT |
| | | right | GTTTTACATTCAACTCAT |
| | S:483del | left | pTAAAGGTCCTAATTGTTAC |
| | | right | TAACAAACCTTGTAAAGG |
| | S:F157S/R158G | left | pGAGTTTATTCTAGTGCGAA |
| | | right | GGATGGAAAGTGAGTCAG |
| | orf1:A7842G | left | pTGTCTAAGTTAACAAAATG |
| | | right | TGTTATTAGCTCTCAGGC |
| | S:69-70del | left | pTAGCATGGAACCAAGTAAC |
| | | right | TACCATTGGTCCCAGAGA |
| PCR primers | N1_F1 | | GACCCCAAAATCAGCGAAAT |
| | N1_R1 | | TCTGGTTACTGCCAGTTGAATCTG |
| | N3_F1 | | GGGAGCCTTGAATACACCAAAA |
| | N3_R1 | | TGTAGCACGATTGCAGCATTG |
| | PMMoV-F | | GAGTGGTTTGACCTTAACGTTTGA |
| | PMMoV-R | | TTGTCGGTTGCAATGCAAGT |
| | RBD_F2 | | CAACTGAAATCTATCAGGCC |
| | RBD_R1 | | GACTTTTTAGGTCCACAAACAG |
| | RBD_F3 | | GCAGATTCATTTGTAATTAGAGG |
| | RBD_R2 | | AAATATCTCTCTCAAAAGGTTTGAG |
| | MD3_F | | TTCTAGAGTTCCTGATCTTCTGGTC |
| | MD3_R | | TACTAGGTTCCATTCTTCAAGGAGC |
| | RBD_R3 | | CAGTTGCTGATTCTCKTCCTG |
| | RBD_F4 | | GAGTCCAACCAACAGAATCTATTG |
| | RBD_R2D | | CTCTCTCAAAAGGTTTGAGHHTAGA |
| | nsp1_F1 | | GTACGGTCGTAGTGGTGAGACAC |
| | nsp1_R1 | | CTTCATAAGGATCAGTGCCAAGCTC |
| | RBD_R4 | | TAGACTCAGTAAGAACACCTGTG |
| | I5T_F1 | | AGAGTATCATGACGTTCGTGTTG |
| | N55S_F1 | | ACCCAATAATACTGCGTCTTGG |
| | N55S_R1 | | TGTTAATTGGAACGCCTTGTCC |
| | Q613_F1 | | AACAAATACTTCTAACCAGGTTGC |
| | Q613_R1 | | CAAGTAGGAGTAAGTTGATCTGC |
| | Q52_F1 | | CACGTGGTGTTTATTACCCTGA |
| | Q52_R1 | | GCATGGAACCAAGTAACATTGG |
| | F157S/R158G_F1 | | CTGTGAATTTCAATTTTGTAATGATCC |
| | F157S/R158G_R1 | | CTGAGAGACATATTCAAAAGTGC |
| | A7842G_F1 | | CATCTTTACTTTGATAAAGCTGGTC |
| | A7842G_R1 | | GAACCTTTAGTGTTATTAGCTCTC |

| | | |
|---|---|---|
| | S:69-70del_R1 | CAAACCTCTTAGTACCATTGGTC |

**Table S2:** List of final primer concentrations in the PCR master mix used from w40/2022 to w15/2023.

| PCR Primer | Final Concentration |
|---|---|
| N1 F | 75 nM |
| N1 R | 100 nM |
| PMMoV-F | 50 nM |
| PMMoV-R | 67 nM |
| RBD_F2 | 150 nM |
| RBD_R1 | 200 nM |
| RBD_F3 | 250 nM |
| RBD_R2 | 300 nM |
| RBD_F4 | 150 nM |
| RBD_R3 | 200 nM |
| nsp1_F2 | 300 nM |
| nsp1_R2 | 400 nM |
| MD3_F | 150 nM |
| MD3_R | 200 nM |

**Table S3:** List of final primer concentrations in the PCR master mix used from w16/2023 to w32/2023.

| PCR Primer | Final Concentration |
|---|---|
| N3_F1 | 75 nM |
| N3_R1 | 100 nM |
| I5T_F1 | 150 nM |
| N1 R | 200 nM |
| PMMoV-F | 50 nM |
| PMMoV-R | 67 nM |
| RBD_F2 | 150 nM |
| RBD_R1 | 150 nM |
| RBD_R4 | 300 nM |
| RBD_F3 | 200 nM |
| RBD_R2 | 300 nM |

| | |
|---|---|
| RBD_F4 | 150 nM |
| RBD_R3 | 200 nM |
| N55S_F1 | 75 nM |
| N55S_R1 | 100 nM |
| Q613_F1 | 150 nM |
| Q613_R1 | 200 nM |
| MD3_F | 150 nM |
| MD3_R | 200 nM |

**Table S4:** List of final primer concentrations in the PCR master mix used from w33/2023 to w35/2023.

| PCR Primer | Final Concentration |
|---|---|
| N3_F1 | 75 nM |
| N3_R1 | 100 nM |
| I5T_F1 | 150 nM |
| N1 R | 200 nM |
| PMMoV-F | 50 nM |
| PMMoV-R | 67 nM |
| RBD_F2 | 150 nM |
| RBD_R1 | 150 nM |
| RBD_R4 | 300 nM |
| RBD_F3 | 200 nM |
| RBD_R2 | 300 nM |
| RBD_F4 | 150 nM |
| RBD_R3 | 200 nM |
| N55S_F1 | 75 nM |
| N55S_R1 | 100 nM |
| Q613_F1 | 150 nM |
| Q613_R1 | 200 nM |
| Q52_F1 | 150 nM |
| Q52_R1 | 200 nM |

**Table S5:** List of final primer concentrations in the PCR master mix used from w36/2023 to w52/2023.

| PCR Primer | Final Concentration |
|---|---|
| N3_F1 | 75 nM |
| N3_R1 | 100 nM |
| I5T_F1 | 150 nM |
| N1_R1 | 200 nM |
| PMMoV-F1 | 50 nM |
| PMMoV-R1 | 67 nM |
| RBD_F2 | 150 nM |
| RBD_R1 | 150 nM |
| RBD_R4 | 300 nM |
| RBD_F3 | 200 nM |
| RBD_R2 | 300 nM |
| RBD_F4 | 150 nM |
| RBD_R3 | 200 nM |
| N55S_F1 | 75 nM |
| N55S_R1 | 100 nM |
| Q613_F1 | 150 nM |
| Q613_R1 | 200 nM |
| Q52_F1 | 150 nM |
| 69-70del_R1 | 200 nM |
| F157S/R158G_F1 | 150 nM |
| F157S/R158G_R1 | 200 nM |
| A7842G_F1 | 150 nM |
| A7842G_R1 | 200 nM |

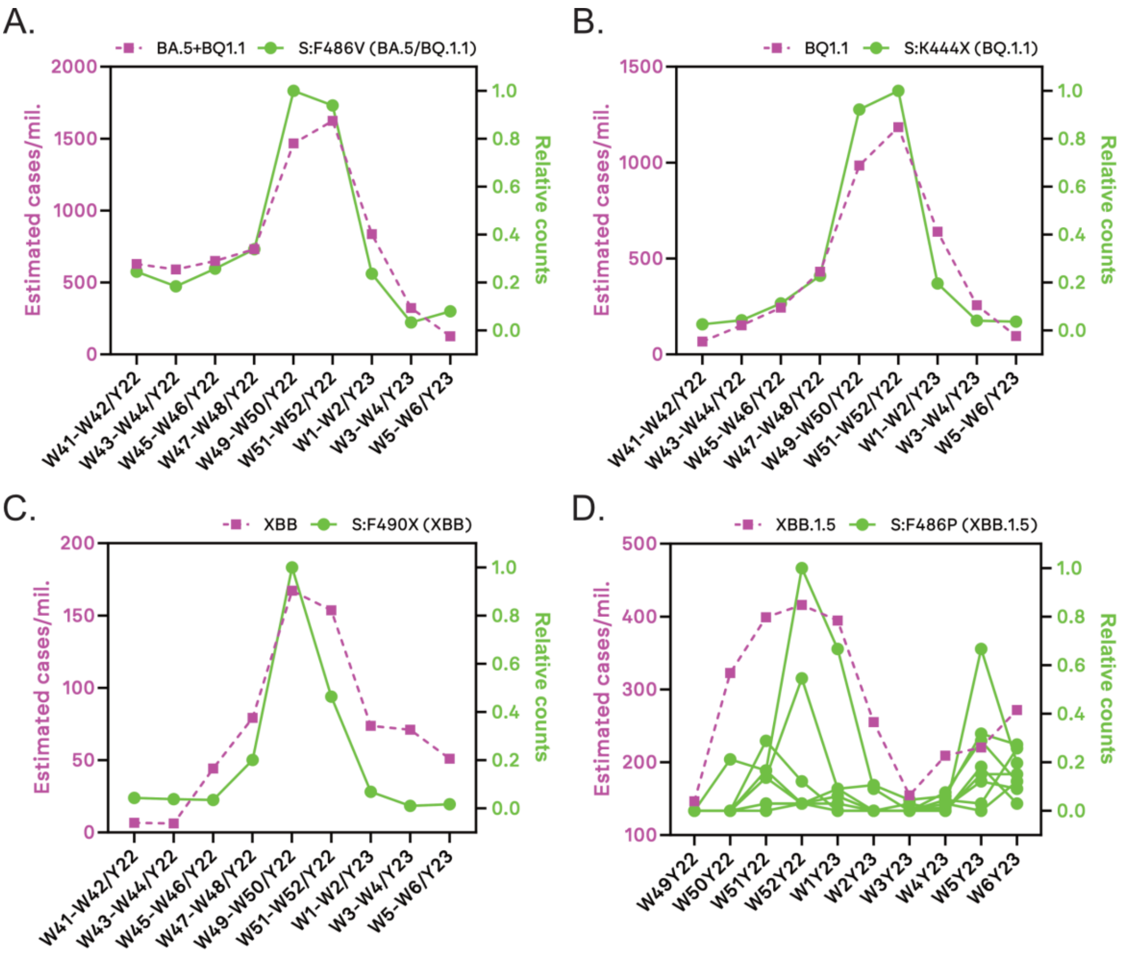


**Figure S1:** Qualitative correlation between observed clinical trends in Sweden for 4 target mutations of concern (square symbols, data manually extracted from GISAID) and relative counts measured with the hpPCR assay (circle symbols). For mutations S:F486V, S:K444X and S:F490X the hpPCR data corresponds to the average of all tested sites, while for S:F486P the results for 8 collection sites corresponding to major populational centers (Helsingborg, Jönköping, Kalmar, Örebro, Östersund, Umeå, Uppsala and Västerås) were individually plotted.

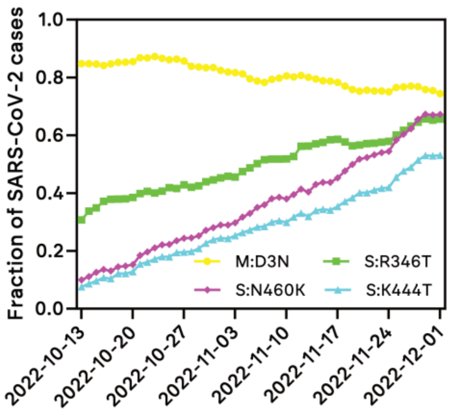

**Figure S2:** Incidence of SARS-CoV-2 cases with each of the 4 target mutations M:D3N, S:N460K, S:R346T and S:K444T between w42/2022 and w48/2022. Data exported from the cov-spectrum.org portal using GISAID data.

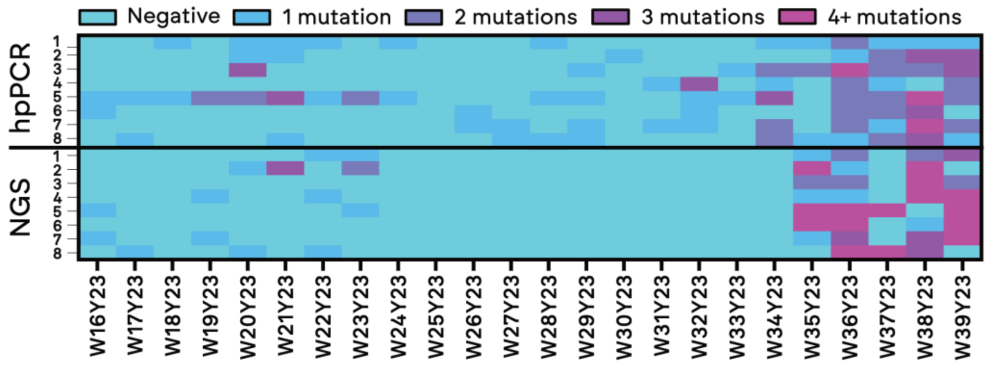


**Figure S3:** Early mutation detection for BA.2.86 using hpPCR (S:483del, S:F157S/R158G, Orf1:A7842G and S:69-70del) and NGS, here considering all mutations characteristic to BA.2.86 captured by whole SARS-CoV-2 genome sequencing.  Details of mutations detected using NGS are listed in Table S6. The selection of markers specific to BA.2.86 was based on a variation of the VaQuERo sensitive marker approach [2].

**Table S6:** List of detected mutations using NGS and respective Allele frequency (AF), quality score (QUAL) and depth (DP) parameters within the timespan shown in Figure S3. AA and NUC refer to the target amino acid/ nucleic acid mutation, respectively.

| Place | year | week | AF | QUAL | DP | AA | NUC |
|---|---|---|---|---|---|---|---|
| Uppsala | 2023 | 16 | 0,026 | 40,6839 | 41807 | M:A104V | C26833T |
| Västerås | 2023 | 17 | 0,0365183 | 111,689 | 27581 | M:A104V | C26833T |
| Örebro | 2023 | 19 | 0,0276382 | 33,5496 | 1194 | M:A104V | C26833T |
| Uppsala | 2023 | 19 | 0,088 | 760,69 | 17038 | ORF1AB:A211D | C897A |
| Helsingborg | 2023 | 20 | 0,0376128 | 114,466 | 12299 | S:S939F | C24378T |
| Västerås | 2023 | 20 | 0,0355533 | 104,416 | 23689 | S:V127F | G21941T |
| Helsingborg | 2023 | 21 | 0,02 | 14,7774 | 39274 | S:Y1215Y | C25207T |
| Helsingborg | 2023 | 21 | 0,052 | 172,132 | 22879 | N:Q229K | C28958A |
| Helsingborg | 2023 | 21 | 0,0365 | 58,4878 | 13832 | ORF1AB:T2676T | C8293T |
| Örebro | 2023 | 22 | 0,123318 | 2293 | 892 | S:R403K | G22770A |
| Göteborg | 2023 | 22 | 0,010859 | 89 | 1013 | S:R403K | G22770A |
| Västerås | 2023 | 22 | 0,099265 | 1034 | 544 | ORF1AB:V3593F | G11042T |
| Göteborg | 2023 | 23 | 0,013126 | 133 | 1219 | S:R403K | G22770A |
| Helsingborg | 2023 | 23 | 0,013793 | 102 | 725 | S:R403K | G22770A |
| Helsingborg | 2023 | 23 | 0,097056 | 4647 | 2514 | M:A104V | C26833T |
| Stockholm-Käppala | 2023 | 23 | 0,314578 | 3106 | 391 | M:A104V | C26833T |
| Västerås | 2023 | 25 | 0,990975 | 18128 | 554 | M:A104V | C26833T |
| Malmö | 2023 | 30 | 0,009842 | 98 | 2337 | M:A104V | C26833T |
| Göteborg | 2023 | 35 | 0,031 | 70,7257 | 21344 | M:A104V | C26833T |
| Helsingborg | 2023 | 35 | 0,0457746 | 123,407 | 21923 | ORF1AB:V3593F | G11042T |
| Helsingborg | 2023 | 35 | 0,0815408 | 658,895 | 22726 | ORF1AB:N4358N | T13339C |
| Helsingborg | 2023 | 35 | 0,0549898 | 101,711 | 981 | ORF1AB:S5164S | T15756A |

| | | | | | | | |
|---|---|---|---|---|---|---|---|
| Helsingborg | 2023 | 35 | 0,357143 | 1044,99 | 287 | S:S50L | C21711T |
| Helsingborg | 2023 | 35 | 0,0425213 | 164,739 | 11704 | S:V127F | G21941T |
| Helsingborg | 2023 | 35 | 0,037 | 115,329 | 76236 | S:A570V | C23271T |
| Helsingborg | 2023 | 35 | 0,029 | 57,7004 | 5169 | M:T30A | A26610G |
| Helsingborg | 2023 | 35 | 0,0855 | 720,923 | 7807 | M:A104V | C26833T |
| Helsingborg | 2023 | 35 | 0,088 | 756,151 | 6369 | ORF1AB:V1056L | G3431T |
| Helsingborg | 2023 | 35 | 0,0415208 | 154,664 | 23119 | ORF1AB:N2526S | A7842G |
| Helsingborg | 2023 | 35 | 0,037 | 63,3236 | 23626 | ORF1AB:T2676T | C8293T |
| Helsingborg | 2023 | 35 | 0,064 | 408,954 | 24920 | ORF1AB:A211D | C897A |
| Malmö | 2023 | 35 | 0,039 | 132,642 | 47174 | S:R158G | A22034G |
| Malmö | 2023 | 35 | 0,0465 | 203,746 | 26513 | M:A104V | C26833T |
| Stockholm-Käppala | 2023 | 35 | 0,0487437 | 212,127 | 54934 | ORF1AB:V3593F | G11042T |
| Stockholm-Käppala | 2023 | 35 | 0,0260521 | 16,1343 | 2073 | ORF1AB:S5164S | T15756A |
| Stockholm-Käppala | 2023 | 35 | 0,0211346 | 19,3826 | 1797 | S:R403K | G22770A |
| Stockholm-Käppala | 2023 | 35 | 0,0341709 | 88,2418 | 20791 | S:S939F | C24378T |
| Stockholm-Käppala | 2023 | 35 | 0,0625313 | 389,029 | 20427 | M:D3H | G26529C |
| Stockholm-Käppala | 2023 | 35 | 0,0445 | 183,659 | 26874 | M:A104V | C26833T |
| Stockholm-Käppala | 2023 | 35 | 0,0425 | 164,125 | 58735 | ORF1AB:N2526S | A7842G |
| Stockholm-Käppala | 2023 | 35 | 0,0515 | 165,261 | 49984 | ORF1AB:T2676T | C8293T |
| Stockholm-Käppala | 2023 | 35 | 0,023023 | 26,3305 | 54893 | ORF1AB:A211D | C897A |
| Umeå | 2023 | 35 | 0,0735586 | 137,874 | 498 | S:R403K | G22770A |
| Umeå | 2023 | 35 | 0,058104 | 114,541 | 636 | S:N481K | T23005A |
| Umeå | 2023 | 35 | 0,072 | 517,91 | 37631 | S:A570V | C23271T |
| Umeå | 2023 | 35 | 0,1555 | 1692,84 | 5364 | S:P1143L | C24990T |
| Umeå | 2023 | 35 | 0,0895 | 783,384 | 27415 | S:Y1215Y | C25207T |
| Umeå | 2023 | 35 | 0,0837139 | 670,632 | 1970 | M:D3H | G26529C |
| Umeå | 2023 | 35 | 0,122122 | 1359,73 | 3532 | M:T30A | A26610G |
| Umeå | 2023 | 35 | 0,074 | 546,929 | 10105 | M:A104V | C26833T |
| Umeå | 2023 | 35 | 0,049 | 146,036 | 19755 | N:Q229K | C28958A |
| Umeå | 2023 | 35 | 0,131 | 1528,44 | 13522 | ORF1AB:N2526S | A7842G |
| Uppsala | 2023 | 35 | 0,0620504 | 206,693 | 1100 | S:R403K | G22770A |
| Örebro | 2023 | 36 | 0,0748031 | 71,3022 | 250 | S:R403K | G22770A |
| Göteborg | 2023 | 36 | 0,418848 | 3503,29 | 744 | S:S50L | C21711T |
| Göteborg | 2023 | 36 | 0,0955 | 884,219 | 27421 | M:A104V | C26833T |
| Helsingborg | 2023 | 36 | 0,043 | 169,139 | 13227 | ORF1AB:N4358N | T13339C |
| Malmö | 2023 | 36 | 0,370253 | 1228,15 | 317 | S:S50L | C21711T |
| Malmö | 2023 | 36 | 0,020572 | 19,6614 | 14348 | S:S939F | C24378T |
| Stockholm-Käppala | 2023 | 36 | 0,0361809 | 59,1855 | 35503 | ORF1AB:V3593F | G11042T |
| Stockholm-Käppala | 2023 | 36 | 0,0245245 | 33,2494 | 28917 | ORF1AB:N4358N | T13339C |
| Stockholm-Käppala | 2023 | 36 | 0,117195 | 658,936 | 1037 | S:S50L | C21711T |
| Stockholm-Käppala | 2023 | 36 | 0,0235235 | 28,8615 | 25175 | S:V127F | G21941T |
| Stockholm-Käppala | 2023 | 36 | 0,0326469 | 86,3826 | 17093 | S:S939F | C24378T |
| Stockholm-Käppala | 2023 | 36 | 0,079 | 461,509 | 19117 | S:P1143L | C24990T |
| Stockholm-Käppala | 2023 | 36 | 0,044 | 177,573 | 52892 | S:Y1215Y | C25207T |
| Stockholm-Käppala | 2023 | 36 | 0,0325325 | 80,6198 | 13395 | M:D3H | G26529C |
| Stockholm-Käppala | 2023 | 36 | 0,038038 | 124,076 | 14209 | M:T30A | A26610G |
| Stockholm-Käppala | 2023 | 36 | 0,0455228 | 193,594 | 17128 | M:A104V | C26833T |
| Stockholm-Käppala | 2023 | 36 | 0,0415208 | 90,7043 | 22632 | N:Q229K | C28958A |
| Umeå | 2023 | 36 | 0,0441545 | 172,733 | 28088 | ORF1AB:V3593F | G11042T |
| Umeå | 2023 | 36 | 0,132066 | 1560,05 | 18868 | ORF1AB:N4358N | T13339C |
| Umeå | 2023 | 36 | 0,0702746 | 220,674 | 1237 | ORF1AB:S5164S | T15756A |
| Umeå | 2023 | 36 | 0,572289 | 3646,5 | 494 | S:S50L | C21711T |
| Umeå | 2023 | 36 | 0,0395198 | 137,551 | 25545 | S:V127F | G21941T |
| Umeå | 2023 | 36 | 0,0435 | 173,595 | 53463 | S:A570V | C23271T |
| Umeå | 2023 | 36 | 0,0285 | 23,5129 | 12448 | S:P1143L | C24990T |
| Umeå | 2023 | 36 | 0,054 | 284,212 | 43054 | S:Y1215Y | C25207T |
| Umeå | 2023 | 36 | 0,048024 | 218,164 | 10163 | M:D3H | G26529C |
| Umeå | 2023 | 36 | 0,0610916 | 372,283 | 8201 | M:T30A | A26610G |
| Umeå | 2023 | 36 | 0,0385 | 128,456 | 19205 | M:A104V | C26833T |

| | | | | | | | |
|---|---|---|---|---|---|---|---|
| Umeå | 2023 | 36 | 0,123 | 1102,82 | 16866 | N:Q229K | C28958A |
| Umeå | 2023 | 36 | 0,108554 | 1108,94 | 9073 | ORF1AB:V1056L | G3431T |
| Umeå | 2023 | 36 | 0,0825 | 670,939 | 19796 | ORF1AB:N2526S | A7842G |
| Umeå | 2023 | 36 | 0,094 | 661,919 | 15991 | ORF1AB:T2676T | C8293T |
| Umeå | 2023 | 36 | 0,127064 | 1458,31 | 32187 | ORF1AB:A211D | C897A |
| Uppsala | 2023 | 36 | 0,0255128 | 38,2542 | 25421 | ORF1AB:N4358N | T13339C |
| Uppsala | 2023 | 36 | 0,0396825 | 50,4595 | 629 | S:R403K | G22770A |
| Uppsala | 2023 | 36 | 0,0205103 | 16,6749 | 8964 | M:A104V | C26833T |
| Västerås | 2023 | 36 | 0,0275 | 48,9472 | 9088 | ORF1AB:N4358N | T13339C |
| Västerås | 2023 | 36 | 0,0628141 | 62,3214 | 398 | ORF1AB:S5164S | T15756A |
| Västerås | 2023 | 36 | 0,648649 | 2334,89 | 259 | S:S50L | C21711T |
| Västerås | 2023 | 36 | 0,049049 | 227,867 | 8232 | S:V127F | G21941T |
| Västerås | 2023 | 36 | 0,031401 | 22,5868 | 404 | S:N481K | T23005A |
| Västerås | 2023 | 36 | 0,0725363 | 526,408 | 26735 | S:A570V | C23271T |
| Västerås | 2023 | 36 | 0,0552486 | 282,044 | 4172 | S:S939F | C24378T |
| Västerås | 2023 | 36 | 0,0425 | 98,5516 | 5282 | S:P1143L | C24990T |
| Västerås | 2023 | 36 | 0,0628242 | 341,238 | 1734 | M:D3H | G26529C |
| Västerås | 2023 | 36 | 0,0389239 | 116,782 | 1750 | M:T30A | A26610G |
| Västerås | 2023 | 36 | 0,0505 | 246,114 | 3579 | M:A104V | C26833T |
| Västerås | 2023 | 36 | 0,0335 | 43,2827 | 5935 | N:Q229K | C28958A |
| Stockholm-Käppala | 2023 | 37 | 0,167894 | 3872 | 1358 | ORF1AB:S5164S | T15756A |
| Stockholm-Käppala | 2023 | 37 | 0,107754 | 7837 | 4733 | S:V127F | G21941T |
| Stockholm-Käppala | 2023 | 37 | 0,062829 | 2885 | 3995 | S:R158G | A22034G |
| Stockholm-Käppala | 2023 | 37 | 0,101607 | 8209 | 5413 | M:A104V | C26833T |
| Stockholm-Käppala | 2023 | 37 | 0,027176 | 397 | 2355 | N:Q229K | C28958A |
| Stockholm-Käppala | 2023 | 37 | 0,2 | 335 | 90 | ORF1AB:V1056L | G3431T |
| Stockholm-Käppala | 2023 | 37 | 0,178734 | 5253 | 2417 | ORF1AB:N2526S | A7842G |
| Västerås | 2023 | 37 | 0,109042 | 6527 | 3705 | ORF1AB:V3593F | G11042T |
| Västerås | 2023 | 37 | 0,205674 | 13414 | 3243 | ORF1AB:N4358N | T13339C |
| Västerås | 2023 | 37 | 0,221311 | 7856 | 1830 | ORF1AB:S5164S | T15756A |
| Västerås | 2023 | 37 | 0,013203 | 77 | 1742 | S:S50L | C21711T |
| Västerås | 2023 | 37 | 0,208802 | 21146 | 4885 | S:V127F | G21941T |
| Västerås | 2023 | 37 | 0,145398 | 9541 | 4161 | S:R158G | A22034G |
| Västerås | 2023 | 37 | 0,183442 | 4506 | 1232 | S:R403K | G22770A |
| Västerås | 2023 | 37 | 0,316501 | 41084 | 5545 | S:A570V | C23271T |
| Västerås | 2023 | 37 | 0,259259 | 1975 | 351 | S:P1143L | C24990T |
| Västerås | 2023 | 37 | 0,045161 | 89 | 310 | S:Y1215Y | C25207T |
| Västerås | 2023 | 37 | 0,137764 | 9898 | 3927 | M:D3H | G26529C |
| Västerås | 2023 | 37 | 0,090414 | 6734 | 4778 | M:T30A | A26610G |
| Västerås | 2023 | 37 | 0,247495 | 15292 | 2994 | N:Q229K | C28958A |
| Västerås | 2023 | 37 | 0,380952 | 388 | 42 | ORF1AB:V1056L | G3431T |
| Västerås | 2023 | 37 | 0,115876 | 2406 | 2192 | ORF1AB:N2526S | A7842G |
| Västerås | 2023 | 37 | 0,21742 | 19291 | 4512 | ORF1AB:T2676T | C8293T |
| Örebro | 2023 | 38 | 0,133054 | 8917 | 3818 | ORF1AB:V3593F | G11042T |
| Örebro | 2023 | 38 | 0,370347 | 14077 | 1612 | ORF1AB:S5164S | T15756A |
| Örebro | 2023 | 38 | 0,065024 | 4940 | 5844 | S:A570V | C23271T |
| Örebro | 2023 | 38 | 0,122271 | 436 | 229 | S:P1143L | C24990T |
| Örebro | 2023 | 38 | 0,040685 | 2007 | 5137 | M:A104V | C26833T |
| Örebro | 2023 | 38 | 0,067829 | 1892 | 2580 | N:Q229K | C28958A |
| Örebro | 2023 | 38 | 0,19437 | 7022 | 2629 | ORF1AB:N2526S | A7842G |
| Örebro | 2023 | 38 | 0,075064 | 4463 | 4303 | ORF1AB:A211D | C897A |
| Göteborg | 2023 | 38 | 0,130114 | 11498 | 4742 | M:D3H | G26529C |
| Göteborg | 2023 | 38 | 0,082643 | 7631 | 6159 | M:T30A | A26610G |
| Helsingborg | 2023 | 38 | 0,243446 | 1361 | 267 | S:S939F | C24378T |
| Helsingborg | 2023 | 38 | 0,156489 | 1345 | 524 | S:P1143L | C24990T |
| Helsingborg | 2023 | 38 | 0,035135 | 95 | 370 | S:Y1215Y | C25207T |
| Helsingborg | 2023 | 38 | 0,060259 | 2876 | 4630 | M:A104V | C26833T |
| Helsingborg | 2023 | 38 | 0,096936 | 2609 | 2187 | N:Q229K | C28958A |
| Helsingborg | 2023 | 38 | 0,105179 | 2516 | 2491 | ORF1AB:N2526S | A7842G |

| | | | | | | | |
|---|---|---|---|---|---|---|---|
| Helsingborg | 2023 | 38 | 0,285714 | 16862 | 2639 | ORF1AB:A211D | C897A |
| Malmö | 2023 | 38 | 0,079793 | 1223 | 965 | S:R403K | G22770A |
| Malmö | 2023 | 38 | 0,016865 | 190 | 2016 | S:N481K | T23005A |
| Malmö | 2023 | 38 | 0,009904 | 76 | 2928 | S:A570V | C23271T |
| Malmö | 2023 | 38 | 0,165803 | 10501 | 3281 | M:D3H | G26529C |
| Malmö | 2023 | 38 | 0,100376 | 6862 | 4254 | M:T30A | A26610G |
| Malmö | 2023 | 38 | 0,035608 | 1403 | 4381 | M:A104V | C26833T |
| Malmö | 2023 | 38 | 0,109009 | 3209 | 2220 | N:Q229K | C28958A |
| Malmö | 2023 | 38 | 0,347826 | 161 | 23 | ORF1AB:V1056L | G3431T |
| Malmö | 2023 | 38 | 0,42933 | 13025 | 1507 | ORF1AB:N2526S | A7842G |
| Malmö | 2023 | 38 | 0,349425 | 28005 | 3480 | ORF1AB:T2676T | C8293T |
| Umeå | 2023 | 38 | 0,16563 | 23032 | 7396 | M:A104V | C26833T |
| Uppsala | 2023 | 38 | 0,163327 | 12097 | 3992 | ORF1AB:N4358N | T13339C |
| Uppsala | 2023 | 38 | 0,190476 | 5719 | 1596 | ORF1AB:S5164S | T15756A |
| Uppsala | 2023 | 38 | 0,064437 | 1140 | 2840 | ORF1AB:N2526S | A7842G |
| Västerås | 2023 | 38 | 0,095814 | 8072 | 5542 | S:A570V | C23271T |
| Västerås | 2023 | 38 | 0,037288 | 1719 | 5015 | M:A104V | C26833T |
| Västerås | 2023 | 38 | 0,062381 | 2145 | 3158 | N:Q229K | C28958A |
| Örebro | 2023 | 39 | 0,1367 | 4784 | 2297 | ORF1AB:S5164S | T15756A |
| Örebro | 2023 | 39 | 0,190539 | 26586 | 6870 | S:V127F | G21941T |
| Örebro | 2023 | 39 | 0,13217 | 11058 | 5561 | S:R158G | A22034G |
| Örebro | 2023 | 39 | 0,053363 | 1582 | 2230 | S:R403K | G22770A |
| Örebro | 2023 | 39 | 0,119988 | 13044 | 6509 | S:A570V | C23271T |
| Örebro | 2023 | 39 | 0,05891 | 434 | 679 | S:S939F | C24378T |
| Örebro | 2023 | 39 | 0,075253 | 6890 | 5940 | M:D3H | G26529C |
| Örebro | 2023 | 39 | 0,049256 | 4267 | 7390 | M:T30A | A26610G |
| Örebro | 2023 | 39 | 0,032781 | 1520 | 5674 | M:A104V | C26833T |
| Örebro | 2023 | 39 | 0,069131 | 3375 | 4166 | N:Q229K | C28958A |
| Örebro | 2023 | 39 | 0,101562 | 330 | 256 | ORF1AB:V1056L | G3431T |
| Örebro | 2023 | 39 | 0,167258 | 16054 | 5076 | ORF1AB:A211D | C897A |
| Göteborg | 2023 | 39 | 0,121557 | 6769 | 3340 | ORF1AB:V3593F | G11042T |
| Göteborg | 2023 | 39 | 0,059051 | 1470 | 2591 | N:Q229K | C28958A |
| Göteborg | 2023 | 39 | 0,095294 | 7110 | 5100 | ORF1AB:T2676T | C8293T |
| Malmö | 2023 | 39 | 0,119957 | 6974 | 3743 | N:Q229K | C28958A |
| Malmö | 2023 | 39 | 0,182109 | 9369 | 3756 | ORF1AB:N2526S | A7842G |
| Stockholm-Käppala | 2023 | 39 | 0,200914 | 20272 | 5032 | ORF1AB:V3593F | G11042T |
| Stockholm-Käppala | 2023 | 39 | 0,153515 | 10146 | 3713 | ORF1AB:N4358N | T13339C |
| Stockholm-Käppala | 2023 | 39 | 0,071169 | 1569 | 1925 | ORF1AB:S5164S | T15756A |
| Stockholm-Käppala | 2023 | 39 | 0,188682 | 27072 | 7086 | S:V127F | G21941T |
| Stockholm-Käppala | 2023 | 39 | 0,126082 | 10667 | 5544 | S:R158G | A22034G |
| Stockholm-Käppala | 2023 | 39 | 0,195309 | 10575 | 2601 | S:R403K | G22770A |
| Stockholm-Käppala | 2023 | 39 | 0,007698 | 97 | 4027 | S:N481K | T23005A |
| Stockholm-Käppala | 2023 | 39 | 0,341426 | 49314 | 6549 | S:A570V | C23271T |
| Stockholm-Käppala | 2023 | 39 | 0,131696 | 1697 | 896 | S:P1143L | C24990T |
| Stockholm-Käppala | 2023 | 39 | 0,332902 | 5684 | 772 | S:Y1215Y | C25207T |
| Stockholm-Käppala | 2023 | 39 | 0,059112 | 4370 | 5295 | M:D3H | G26529C |
| Stockholm-Käppala | 2023 | 39 | 0,0299 | 1812 | 6689 | M:T30A | A26610G |
| Stockholm-Käppala | 2023 | 39 | 0,068757 | 4714 | 5454 | M:A104V | C26833T |
| Stockholm-Käppala | 2023 | 39 | 0,416599 | 37375 | 3687 | N:Q229K | C28958A |
| Stockholm-Käppala | 2023 | 39 | 0,170576 | 1298 | 469 | ORF1AB:V1056L | G3431T |
| Stockholm-Käppala | 2023 | 39 | 0,224234 | 9649 | 3037 | ORF1AB:N2526S | A7842G |
| Stockholm-Käppala | 2023 | 39 | 0,215396 | 18550 | 4183 | ORF1AB:A211D | C897A |
| Umeå | 2023 | 39 | 0,366018 | 48983 | 5497 | ORF1AB:V3593F | G11042T |
| Umeå | 2023 | 39 | 0,214787 | 23230 | 5126 | ORF1AB:N4358N | T13339C |
| Umeå | 2023 | 39 | 0,288946 | 19500 | 3094 | ORF1AB:S5164S | T15756A |
| Umeå | 2023 | 39 | 0,161315 | 24233 | 7817 | S:V127F | G21941T |
| Umeå | 2023 | 39 | 0,111571 | 10444 | 6283 | S:R158G | A22034G |
| Umeå | 2023 | 39 | 0,210634 | 12981 | 2934 | S:R403K | G22770A |
| Umeå | 2023 | 39 | 0,288594 | 49314 | 7987 | S:A570V | C23271T |

| | | | | | | | |
|---|---|---|---|---|---|---|---|
| Umeå | 2023 | 39 | 0,301136 | 4827 | 704 | S:S939F | C24378T |
| Umeå | 2023 | 39 | 0,292008 | 5805 | 976 | S:P1143L | C24990T |
| Umeå | 2023 | 39 | 0,094303 | 505 | 509 | S:Y1215Y | C25207T |
| Umeå | 2023 | 39 | 0,342064 | 49314 | 6414 | M:D3H | G26529C |
| Umeå | 2023 | 39 | 0,239911 | 43199 | 8053 | M:T30A | A26610G |
| Umeå | 2023 | 39 | 0,128361 | 14165 | 6435 | M:A104V | C26833T |
| Umeå | 2023 | 39 | 0,062937 | 365 | 572 | ORF1AB:V1056L | G3431T |
| Umeå | 2023 | 39 | 0,422709 | 46805 | 5020 | ORF1AB:N2526S | A7842G |
| Umeå | 2023 | 39 | 0,281698 | 43307 | 6830 | ORF1AB:T2676T | C8293T |
| Umeå | 2023 | 39 | 0,097867 | 8942 | 5814 | ORF1AB:A211D | C897A |
| Uppsala | 2023 | 39 | 0,405518 | 11441 | 1196 | ORF1AB:S5164S | T15756A |
| Uppsala | 2023 | 39 | 0,11288 | 9191 | 4961 | S:A570V | C23271T |
| Uppsala | 2023 | 39 | 0,151976 | 859 | 329 | S:S939F | C24378T |
| Uppsala | 2023 | 39 | 0,101446 | 7791 | 4426 | M:D3H | G26529C |
| Uppsala | 2023 | 39 | 0,067624 | 4822 | 5294 | M:T30A | A26610G |
| Uppsala | 2023 | 39 | 0,132194 | 8434 | 3979 | M:A104V | C26833T |
| Uppsala | 2023 | 39 | 0,138549 | 5003 | 2288 | N:Q229K | C28958A |
| Uppsala | 2023 | 39 | 0,244438 | 18787 | 3596 | ORF1AB:A211D | C897A |

**Section S1**: ddPCR was performed using a One-step RT-ddPCR Advanced Kit for Probes and a Bio-Rad QX600 AutoDG Droplet Digital System according to the manufacturers instructions. Briefly, 4 uL of each wastewater sample were combined with Supermix, reverse transcriptase, 300 mM DTT and primer sequences to a final volume of 20 µL. Droplets were then generated in an automated droplet station (AutoDG instrument), followed by the following temperature cycling protocol in a Bio-rad C1000 deep-well thermocycler instrument: 50 °C for 60 min, 95 °C for 10 min, 40x cycles of (95 °C for 30 s, 60 °C for 60 s), 98 °C for 10 min and finally 4 °C for 30 min. After amplification, the droplets were measured in a QX600 instrument.

**Table S7:** Details of each of the 22 selected wastewater sampling sites

| Areas | WWTP | Served population | Start of sampling period | End of sampling period | Comment |
|---|---|---|---|---|---|
| Gävle | Duvbackens avloppsreningsverk | 89 000 | w46/2022 | | |
| Göteborg | Ryaverket | 800 000 | w03/2023 | | |
| Helsingborg | Öresundsverket | 149 000 | w43/2022 | | |
| Jönköping | Simsholmens avloppsreningsverk | 71 000 | w42/2022 | | |
| Kalmar | Kalmarsundsverket | 66 000 | w40/2022 | | |
| Karlstad | Sjöstadsverket | 70 000 | w21/2023 | | |
| Linköping | Nykvarnsverket | 154 000 | w22/2023 | | |
| Luleå | Uddebo reningsverk | 70 000 | w21/2023 | | |
| Malmö | Sjölunda reningsverk | 356 000 | w03/2023 | | |
| Örebro | Skebäcks reningsverk | 137 000 | w40/2022 | | |
| Östersund | Gövikens reningsverk | 54 000 | w43/2022 | | |
| Stockholm-Bromma | Bromma reningsverk | 375 000 | w25/2023 | | Flow-compensated mix of inlets |

| | | | | | |
|---|---|---|---|---|---|
| Stockholm-Grödinge | Himmerfjärdsverket | 345 000 | w22/2023 | | |
| Stockholm-Hässelby | Bromma reningsverk | | w40/2022 | w22/2023 | Inlet of Stockholm-Bromma |
| Stockholm-Henriksdal | Henriksdals reningsverk | 875 000 | w27/2023 | | Flow-compensated mix of inlets |
| Stockholm-Järva | Bromma reningsverk | | w40/2022 | w22/2023 | Inlet of Stockholm-Bromma |
| Stockholm-Käppala | Käppalaverket | 500 000 | w04/2023 | | |
| Stockholm-Riksby | Henriksdals reningsverk | | w40/2022 | w22/2023 | Inlet of Stockholm-Henriksdal |
| Stockholm-Sickla | Bromma reningsverk | | w40/2022 | w22/2023 | Inlet of Stockholm-Bromma |
| Umeå | Öns reningsverk | 109 000 | w40/2022 | | |
| Uppsala | Kungsängsverket | 191 000 | w40/2022 | | |
| Västerås | Kungsängsverket | 145 000 | w43/2022 | | |
